# Design of the COMEBACK and BACKHOME Studies, Longitudinal Cohorts for Comprehensive Deep Phenotyping of Adults with Chronic Low-Back Pain (cLBP): a part of the BACPAC Research Program

**DOI:** 10.1101/2024.04.09.24305574

**Authors:** Trisha F. Hue, Jeffrey C. Lotz, Patricia Zheng, Dennis M. Black, Jeannie Bailey, Susan K. Ewing, Aaron J. Fields, Wolf Mehling, Aaron Scheffler, Irina Strigo, Thomas Petterson, Lucy A. Wu, Conor O’Neill, The UCSF REACH Center, the CoRe CEnter for PAtient-Centric Mechanistic PHenotyping in Chronic Low Back Pain

## Abstract

**Objective:** The University of California, San Francisco (UCSF) Core Center for Patient-centric, Mechanistic Phenotyping in Chronic Low Back Pain (REACH) is one of the three NIH Back Pain Consortium (BACPAC) Research Programs Mechanistic Research Centers (MRCs). The goal of UCSF REACH is to define cLBP phenotypes and pain mechanisms that can lead to effective, personalized treatments for patients across the population. The primary objective of this research project is to address the critical need for new diagnostic and prognostic markers, and associated patient classification protocols for chronic low back pain (cLBP) treatment.

**Design:** To meet this objective, REACH is conducting two large investigator-initiated translational research cohort studies called: The Longitudinal Clinical Cohort for Comprehensive Deep Phenotyping of Chronic Low-Back Pain (cLBP) Adults Study (comeBACK) and the Chronic Low-Back Pain (cLBP) in Adults Study (BACKHOME).

**Setting:** comeBACK is a longitudinal multicenter in-person observational study of 450 adults with chronic low back pain designed to perform comprehensive deep phenotyping. While, the BACKHOME study is a site-less longitudinal observational e-cohort of approximately 3000 U.S. adults with cLBP. To our knowledge, BACKHOME is the largest prospective remote registry of nationwide adults with cLBP.

**Methods:** Both the comeBACK and BACKHOME studies are collecting a robust and comprehensive set of risk factors, outcomes, and covariates in order to perform deep phenotyping of cLBP patients based on combined biopsychosocial variables to: define cLBP subtypes, establish phenotyping tools for routine clinical evaluation, and lead to improved cLBP outcomes in the future. The data from both studies will be used to establish techniques to develop a patient-centric definition of treatment success and to analyze cLBP patient traits to define clinically useful cLBP phenotypes, using a combination of traditional data analyses and deep learning methods.

**Conclusions:** These 2 pivotal studies, in conjunction with the ancillary studies being performed in both comeBACK and BACKHOME, and the other BACPAC-consortium research projects, we will be able to address a number of diagnostic and therapeutic issues in this complex and diverse patient population with cLBP. These studies will help clarify biopsychosocial mechanisms of cLBP with the aim to provide a foundation to improve the evaluation of treatment effectiveness and to spur new avenues of therapeutic research, including personalized outcome measures that constitute a clinically meaningful treatment effect for individual cLBP patients.

## BACKGROUND

Chronic low back pain (cLBP) is one of the most common forms of pain among adults worldwide^1^ and one of the leading causes of “years lived with disability”^2^. Global studies have found that the annual prevalence of cLPB ranges from 15% to 45%^3^. As a result, cLBP is one of the main reasons for seeking health care services^4–5^, medical costs, sick leave, and individual suffering^5–7^.

Current cLBP treatments are often ineffective, which has led to an increased use of opioids^8^. Biological, psychological, and social factors clearly affect the onset and trajectory of back pain. However, to date, treatments based on the biopsychosocial model have, at best, moderate treatment effects^9^. This is not surprising, as the diagnostic label ‘cLBP’ encompasses many different disease variants and associated patient-specific pathophysiological mechanisms. Consequently, there is an urgent need for new tools to identify clinically-relevant cLBP subtypes and optimize treatment outcomes.

The Back Pain Consortium (BACPAC) Research Program is a component of the Helping to End Addiction Long-term^SM^ Initiative (NIH HEAL Initiative^SM,^ a trans-agency effort to speed scientific solutions to stem the national opioid public health crisis). The goal of BACPAC is to address the need for effective and personalized therapies for cLBP. It will examine biomedical mechanisms within a biopsychosocial context by using interdisciplinary methods and exploring innovative technologies. To accomplish this, BACPAC investigators are characterizing people with cLBP using novel deep phenotyping methodologies to: 1) improve understanding of the complex mechanisms underlying the condition; 2) identify novel pathways and targets for intervention; 3) develop new therapeutic options to reduce pain and improve function; and 4) develop precise diagnostic and treatment algorithms so health care providers can tailor therapies to patients^10^.

The University of California San Francisco’s (UCSF) Core Center for Patient-centric, Mechanistic Phenotyping in Chronic Low Back Pain (REACH) is one of three BACPAC Mechanistic Research Centers (MRCs) charged to recruit and deeply phenotype cLBP patients. The UCSF REACH investigators are conducting translational and clinical research to clarify biopsychosocial mechanisms of chronic low back pain – the interconnection between biology, biomechanics, psychology, and socio-environmental factors – which will be foundational for new diagnostic and therapeutic strategies.

## RATIONALE FOR STUDIES

The goal of UCSF REACH is to define cLBP phenotypes and pain mechanisms that can lead to effective, personalized treatments for patients across the population. The primary objective of this research project is to address the critical need for new diagnostic and prognostic markers, and associated patient classification protocols for cLBP treatment. To meet this objective, REACH is conducting two investigator-initiated studies, comeBACK and BACKHOME, with the aim to determine personalized outcome measures that constitute a clinically meaningful treatment effect for individual patients.

The objectives of both the comeBACK and BACKHOME studies are to collect a robust and comprehensive set of risk factors, outcomes, and covariates in order to perform deep phenotyping of cLBP patients based on combined biopsychosocial variables to: define cLBP subtypes, establish phenotyping tools for routine clinical evaluation, and lead to improved cLBP outcomes in the future. To meet these objectives, we are performing both a traditional longitudinal multicenter cohort study with in-person measurements (comeBACK), as well as a large longitudinal nation-wide remote e-cohort (BACKHOME). Our aim is to use data from both studies to establish techniques to develop a patient-centric definition of treatment success and to analyze cLBP patient traits to define clinically useful cLBP phenotypes, using a combination of traditional data analyses and deep learning methods. Our goal is to provide a foundation to improve the evaluation of treatment effectiveness and to spur new avenues of therapeutic research for chronic low back pain.

### STUDY DESIGNS: comeBACK and BACKHOME

#### comeBACK STUDY

The Longitudinal Clinical Cohort for Comprehensive Deep Phenotyping of Chronic Low-Back Pain (cLBP) Adults Study (comeBACK) is a longitudinal cohort of adults with chronic low back pain. This multicenter observational study was designed to perform comprehensive deep phenotyping in patients with cLBP and is being conducted at 4 clinical sites in the United States (U.S.) with a coordinating center at UCSF. The comeBACK clinical sites are located at four of the University of California campuses, including UC San Francisco (UCSF), UC Davis, UC Irvine, and UC San Diego. These sites within the University of California system were chosen to include Principal Investigators with known expertise in this field, an established clinical site with a wide recruitment pool, as well as to facilitate access to UC-wide electronic health record data for consenting participants.

The original study goal was to enroll 400 men and women. Recruitment commenced in March 2021 and was completed in June 2023 with a total of 450 participants enrolled (113% of goal) and to be followed for up to 2 years. As shown in Figure 1, comeBACK study participants were enrolled and consented at the clinical sites at baseline. Participants attend in-clinic baseline and annual visits (Month 12 and 24). Remote (via online surveys with link sent by email and/or phone) visits occur at Months 1, 2, 3, 4, 5, 6, and 18. Table 2 outlines the measures to be conducted at each of the study visits. Clinical sites made every possible effort to ensure that each study visit is conducted within the specified visit window.

**Figure 1.**
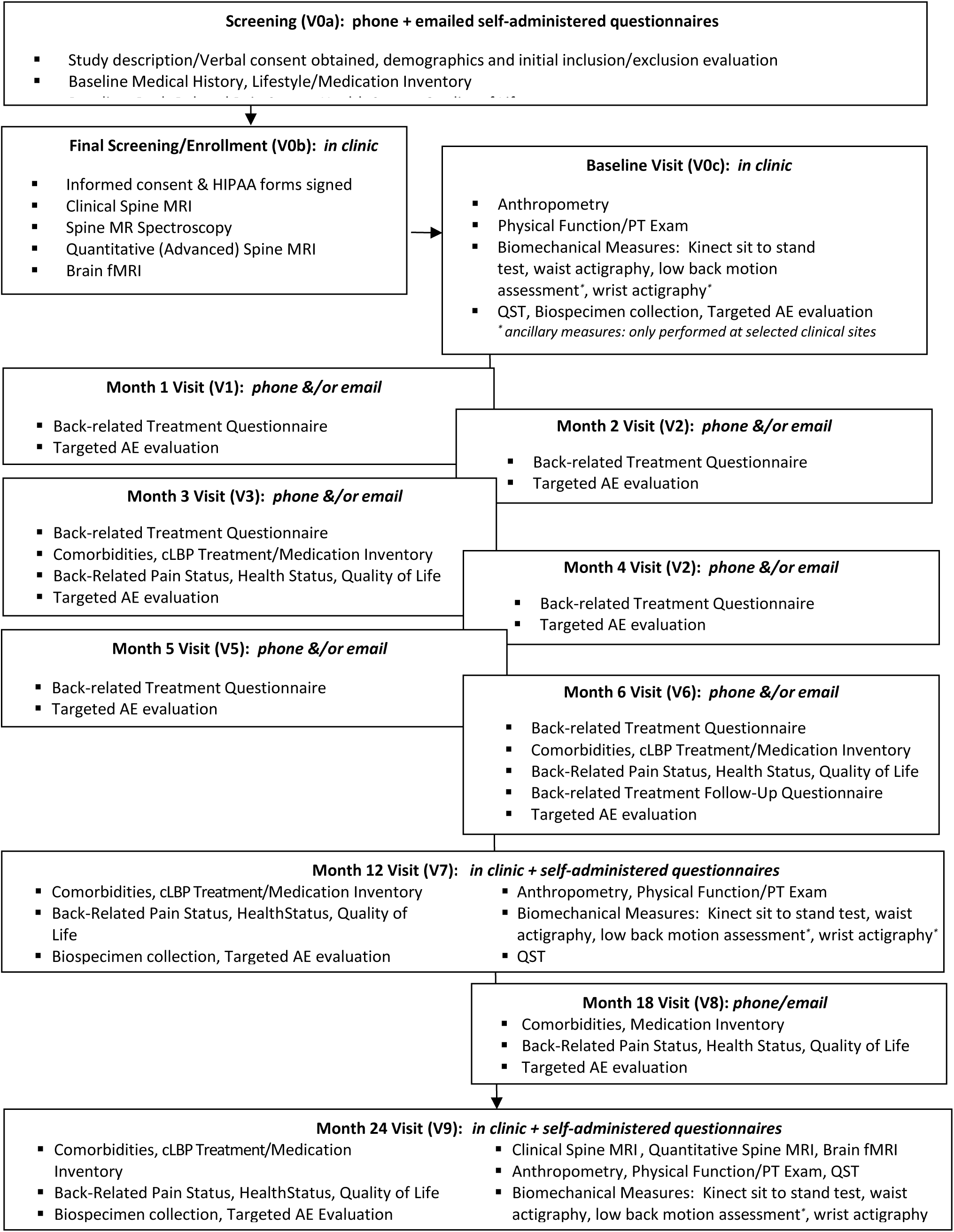
comeBACK Study Flow Diagram

Study governance and advisory boards include: (1) the UCSF Coordinating Center (UCSF CC), which has primary responsibility for the conduct and administration of the study protocol, including management of overall study activities and direction of study partners; development and housing the central study database; development of primary study documentation/Standard Operating Procedures (SOPs), data collection tools, and informed consent; maintaining IRB approval; reporting to the Steering Committee and Observational Study Monitoring Board (OSMB), statistical analysis, receipt of study data from all data sources and archival of data obtained pursuant to the requirements of the study protocol during the course of the study and after the study has been completed. The UCSF CC will also coordinate post-processing analyses with the REACH cores, e.g., Physical Function/Biomechanical Core, Pathophysiology/Imaging Core, etc.; (2) a Steering Committee, consisting of experts in their fields and members from the comeBACK participating units (coordinating center and clinical site Principal Investigators (PIs)), who have responsibility for the scientific direction of the study; (3) an independent Observational Study Monitoring Board (OSMB), who is responsible for safeguarding the interests of study participants, assessing the safety and validity of study procedures, and for monitoring the overall conduct of the study and outcomes data. The OSMB members are independent consultants to the National Institutes of Health-National Institute of arthritis, musculoskeletal and skin diseases (NIH-NIAMS) and are required to provide recommendations about the study. In addition, the OSMB is asked to make recommendations, as appropriate, to the PI about the effects of the study design, benefit/risk ratio of procedures and participant burden; selection, recruitment, and retention of participants; adherence to protocol requirements; completeness, quality, and analysis of measurements; amendments to the studyprotocol; adequacy of and amendments to consent forms; performance of the clinical centers; and participant safety.; and (4) an independent Patient Advisory Boad (PAB), who is responsible for helping to represent the interests of patients and provide feedback on the study.

### BACKHOME STUDY

The online cohort of Chronic Low-Back Pain (cLBP) in Adults (BACKHOME) study is a site-less longitudinal observational e-cohort of U.S. adults with cLBP. Approximately 3000 participants will be enrolled and followed for two years or more. Recruitment commenced in July 2021 and is currently ongoing. To our knowledge, this is the largest prospective registry of nationwide adults with chronic low back pain.

The entire BACKHOME study is conducted remotely and participants are only required to have access to an internet-enabled smartphone, tablet, or computer, to be able to complete the study measures. Participants may choose to either use a website-platform to enroll (using a computer, tablet, or smartphone) or the Eureka mobile application, which can be downloaded onto any smart (iOS or android) phone, to complete the study surveys. There are no in-person visits. See figure 2 for the visit flow and table 3 for the full visit schedule.

**Figure 2:**
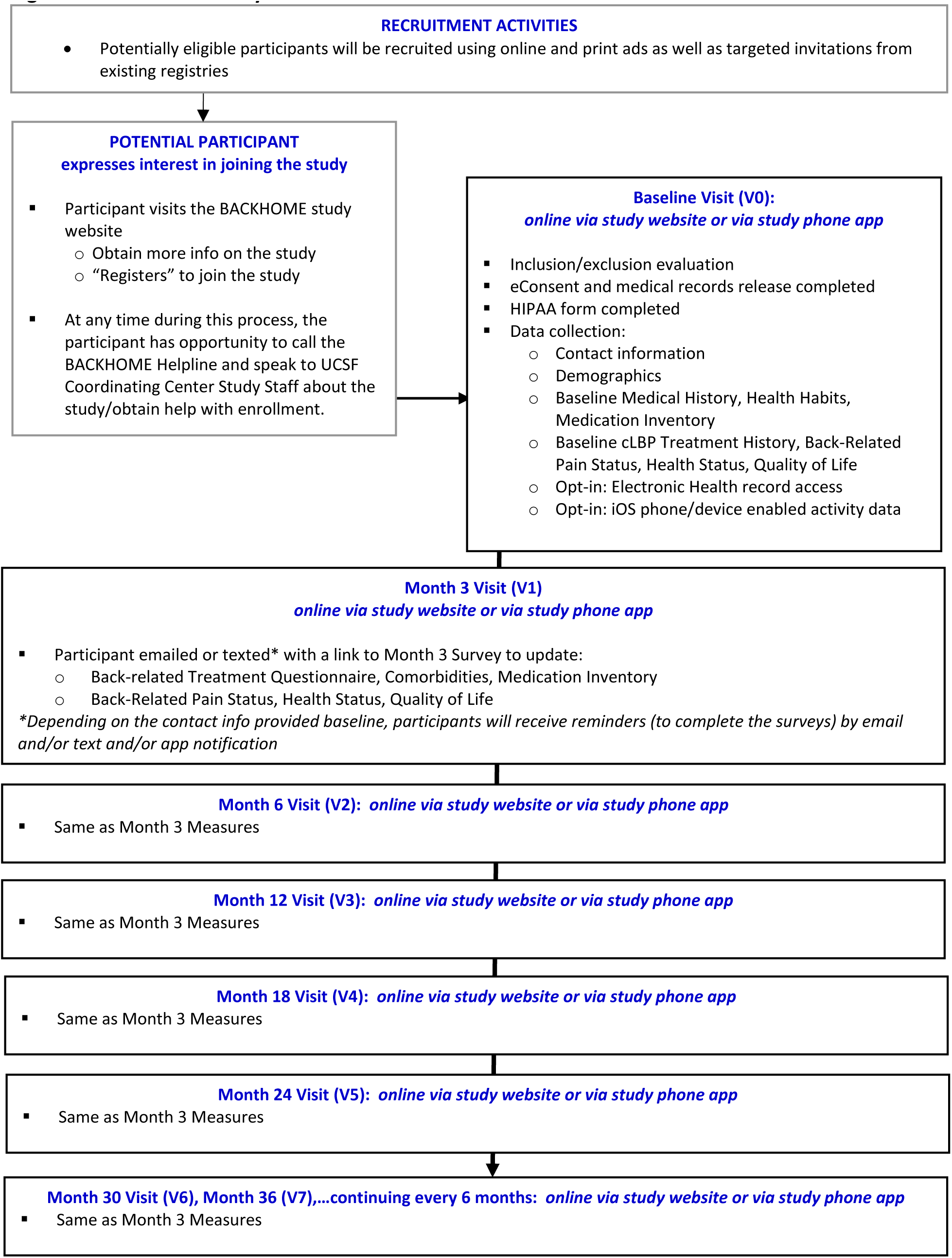
BACKHOME Study Flow

Individuals interested in participating in the study are invited to visit the study website to learn more. If potential participants choose to “join the study”, they initiate enrollment via the study website and are first directed to “register” for an account on the Eureka mobile health platform, an NIH-funded web and mobile app research platform developed at UCSF (http://info.eurekaplatform.org/). Individuals will then answer a few short questions to confirm eligibility, and if eligible, they will be guided through the electronic consent (eConsent) process. The participants will be able to print a copy of the consent from the website for their records and will also be asked to complete the HIPAA authorization form and a medical records release form. At any time during this process, the potential participant has the opportunity to contact the BACKHOME Helpline and speak to UCSF Coordinating Center Study Staff about the study, ask questions, and/or obtain help with enrollment. Participants may also email the UCSF Coordinating Center, if they’d prefer.

After the eConsent has been recorded, the participant will be led through the baseline survey on the website. The participant may choose to complete the questionnaires all at one time, but may take a break at any time and log back into the study website to complete the baseline survey. At any time during the study, a participant may contact the BACKHOME Helpline to speak to UCSF Coordinating Center Study Staff. All participants will be contacted by email, text, and/or push notifications (on their smartphones) for completion of follow-up surveys at Months 3, 6, 12, 18, 24, and will continue approximately every 6 months (until the end of the study). Ascertainment of events may also be obtained through a query of the participant’s electronic medical records (for example: patients in the University of California health systems). Participants can also voluntarily provide permission to collect additional iOS HealthKit data from their smartphones/watches (providing this data is optional and does not otherwise preclude study participation). Vital status/dates and cause of death may also be obtained later using medical/outside records/death registries (such as a National Death Index (NDI) search), if necessary.

Study governance and advisory boards include: (1) the UCSF Coordinating Center will have primary responsibility for conduct/administration of the study protocol, including: management of overall study activities and direction of study partners; development and housing the central study database; development of primary study documentation/SOPs, data collection tools, and informed consent; maintaining IRB approval; reporting to the Steering Committee; statistical analysis; receipt of study data from all data sources and archival of data obtained pursuant to the requirements of the study protocol during the course of the study and after the study has been completed. The UCSF CC will also coordinate post-processing analyses with the REACH cores, e.g., Physical Function/Biomechanical Core, Pathophysiology/Imaging Core, etc.; (2) a Steering Committee, consisting of experts in their fields and members from the comeBACK participating units (coordinating center and clinical site Principal Investigators (PIs)), who has responsibility for the scientific direction of the study; and (3) an independent PAB, who is be responsible for helping to represent the interests of patients and provide feedback on the study.

ETHICAL REVIEW: Both the comeBACK and BACKHOME studies have been approved by the WCG Institutional Review Board (IRB) under reliance agreements from the UCSF, UCD, UCI, and UCSD IRBs.

## STUDY POPULATIONS

The target population for both comeBACK and BACKHOME are adults in the U.S. with chronic low back pain and population-based recruitment methods were employed for both studies in an effort to reach a varied study base. Recruitment approaches were all IRB-approved and included direct mailings, personal and phone invitations, advertisement via various sources, including social media, and referral/word-of mouth.

comeBACK: Potentially eligible participants were recruited at each of the 4 UC clinical sites using a number of methods including: (1) Electronic health record chart review: Individuals were identified via ICD-9/10 code (for study inclusion/exclusion criteria) search of the local site’s electronic medical record system. These potentially eligible patients were contacted by phone or recruitment mailings via: (1) hardcopy mail and (2) MyChart, an online health management system for patients if available at the local clinical site. Interested patients were directed to the study website to complete an online “lead form” or patients could also email or call the local clinical site.; (2) MD-referral method: Patients attending upcoming medical visits at the clinical site (e.g., at the UCSF spine center) were identified and invited to participate. New staff were brought on to review the weekly physician (e.g., at the UCSF spine center) schedules and identify potentially eligible participants. The physicians were then notified weekly with this “eligible” patient list. Either at the clinic visit or via a phone contact, the physician then approached the patient to tell them about the study and determine initial interest. If the patient noted that they would like to learn more, the physician then messaged the Study Coordinator, who followed-up with the participant to continue screening.; (3) Flyers with QR codes (with a link to the study website and/or site contact information), brochures, and a recruitment video were developed by centrally by the Coordinating Center and made available to all clinical sites. Each site developed a local plan for recruitment, and most displayed these tools in UC medical clinics near the recruiting site (e.g., in the reception area of general practitioner physicians’ offices).

BACKHOME: All of the comeBACK recruitment strategies (noted above) are also being used in BACKHOME. Additionally, a number of other recruitment efforts, via convenience sampling, were launched to enroll participants across the U.S., including: (1) Social media ads (e.g., Facebook, Google): Additional targeted recruitment efforts (e.g., ads with visuals including diverse representation and distribution of ads to specific zip codes with racial and income diverse populations) via social media advertisements were also used to reach and increase recruitment of underrepresented populations, including black and indigenous, Latine, Asian, and other diverse communities in the U.S.; (2) Email invitations and notices in newsletters to participants in other research registries (e.g., Eureka Research Platform Participants).

STUDY ELIGIBILITY: The primary eligibility requirement for all BACPAC studies, including comeBACK and BACKHOME, was current chronic low back pain (cLBP) as defined by the NIH Task Force on Research Standards for cLBP^10–11^. The cLBP definition is pain between the lower posterior margin of the rib cage and the horizontal gluteal fold, which has persisted for at least the past 3 months and has resulted in pain on at least 50% of days in the past 6 months. Participant eligibility was primarily determined by self-report during screening. comeBACK eligibility criteria (Table 1) were developed to enroll non-specific cLBP patients, while BACKHOME eligibility allowed for enrollment of a more general cLBP population. Participants were excluded from both studies if they had an inability to read and write in the English language. This was required to help ensure that participants were provided with adequate informed consent, to standardize measurement administration, and also ensure that participants were able to complete the study questionnaires, as intended. Many of the BACPAC minimum dataset questionnaires were not validated in a large variety of languages, so we would not have been able to provide validated versions of the entire minimum dataset to non-English language speakers.

**Table 1:** comeBACK and BACKHOME Eligibility Criteria.

|  | **comeBACK**<br>**Study** | **BACKHOME**<br>**Study** |
| --- | --- | --- |
|  | In-Person<br>Clinical Cohort | eCohort |
| **INCLUSION CRITERIA** | Participants must meet **all** of the following at baseline to eligible: |  |
| Women and men aged $\geq 18$ years | x | x |
| Current self-report of chronic low back pain (pain between the lower posterior margin of the rib cage and the horizontal gluteal fold), which has persisted for ≥ the past 3 months AND has resulted in pain on $\geq 50\%$ of days in the past 6 months* | x | x |
| Current location of worst bodily pain is in the lower back (as compared to pain in other parts of the body) | x |  |
| Life expectancy of at least 2 years | x |  |
| Willing and able to return and comply with scheduled follow up visits (up to $\geq 24$ months of follow-up) | x | |
| Current patient at one of the participating University of California (UC) medical providers/centers | x |  |
| Currently living in the United States |  | x |
| Have access to the internet, email, and a personal mobile smart phone, tablet, or a computer to complete the study measures |  | x |
| Willing and able to provide e-consent | x | x |
| **Exclusion Criteria** | Individuals are excluded at baseline if they meet **any** of the following: |  |
| Any contraindication to MRI** (as defined by the institution performing the MRI), including any of the following: metal in parts of body that will cause safety issues, BMI $\geq 40$, or claustrophobia | x | |
| History of discitis osteomyelitis (spine infection) or spine tumor | x |  |
| History of ankylosing spondylitis, rheumatoid arthritis, polymyalgia rheumatica, psoriatic arthritis, or lupus | x |  |
| History of any bone-related cancer or cancer that metastasized to the bone | x |  |
| Have cancer that metastasized or spread to bones |  |  |
| Currently in treatment for any cancer or plan to start cancer treatment in the next 12 months | x |  |
| History of any cancer treatment in the last 24 months | x |  |
| Had cancer treatment in the last 24 months or currently being treated/will be treated for cancer in next 12 months |  |  |
| Diagnosis of any vertebral fracture in the last 6 months | x |  |
| History of cauda equina syndrome or severe leg weakness (e.g., footdrop) due to your low back problem | x |  |
| Currently diagnosed with cauda equina syndrome or severe leg weakness due to lower back pain |  |  |
| Plan to move out of the area within the next 2 years | x |  |
| Current pregnancy or plan/desire to become pregnant in the next 2 years | x |  |
| Current pregnancy |  |  |
| Inability to read and write in the English language | x | x |
| Current/planned (in the next 2 years) enrollment in another study of a device or investigational drug that would interfere with this study, this may include participation in a blinded trial | x |  |
| Any other criteria, which would make the participant unsuitable to participate in this study as determined by the clinical site Principal Investigator (e.g., an uncontrolled drug and/or alcohol addiction) | x |  |

*\*cLBP criteria as defined by the NIH Pain Consortium Research Task Force (RTF) and BACPAC Minimum Dataset Working Group*
*\*\*Some participants were unable to have ALL of the study MRI imaging scans, but were still eligible as per protocol, e.g., if a participant had MRI safe hardware/artifact between the T12 and S1 vertebrae, the spine MRIs may not have been performed as they would be “unreadable”. Enrollment waivers were provided to participants on a case-by-cases basis after review by the UCSF Coordinating Center.*

## STUDY MEASUREMENTS

The comeBACK study included collection of the entire BACPAC minimum dataset^10^ (including required demographic and outcome measures) and additional BACPAC broadly collected patient reported outcome (PRO) measures^10^, including biomechanical, physical function, physical exam, imaging, and Quantitative Sensory Testing (QST) data. The measures include tracking of changes in risk factors, back-related pain outcomes, and ascertainment of new treatment and medication usage. As BACKHOME had a remote study design, efforts were made to collect as many of the datapoints in the BACPAC minimum dataset, as possible. Tables 2 and 3 outline comeBACK and BACKHOME study visit schedules and the measurements conducted at each visit.

**Table 2:**
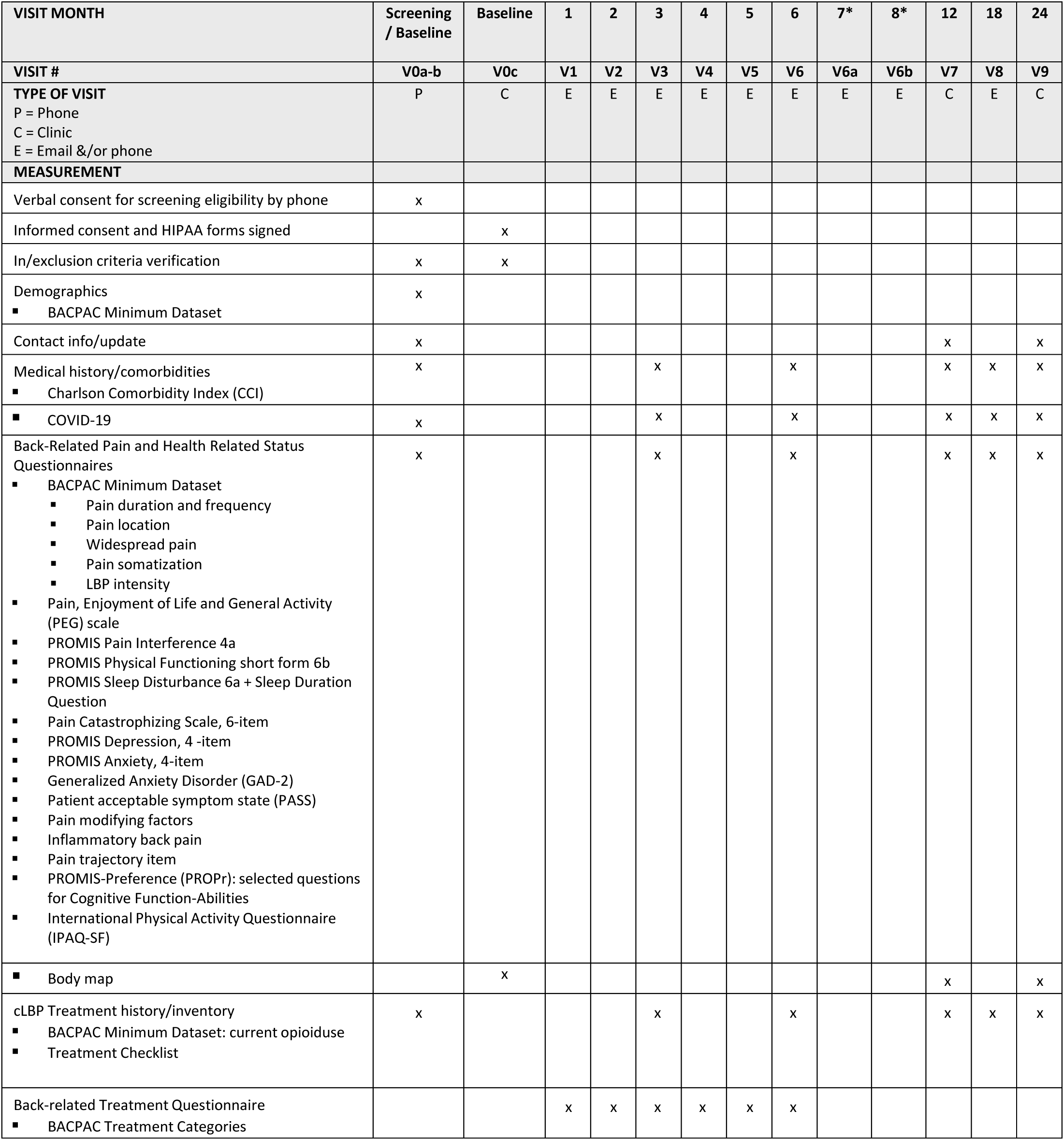
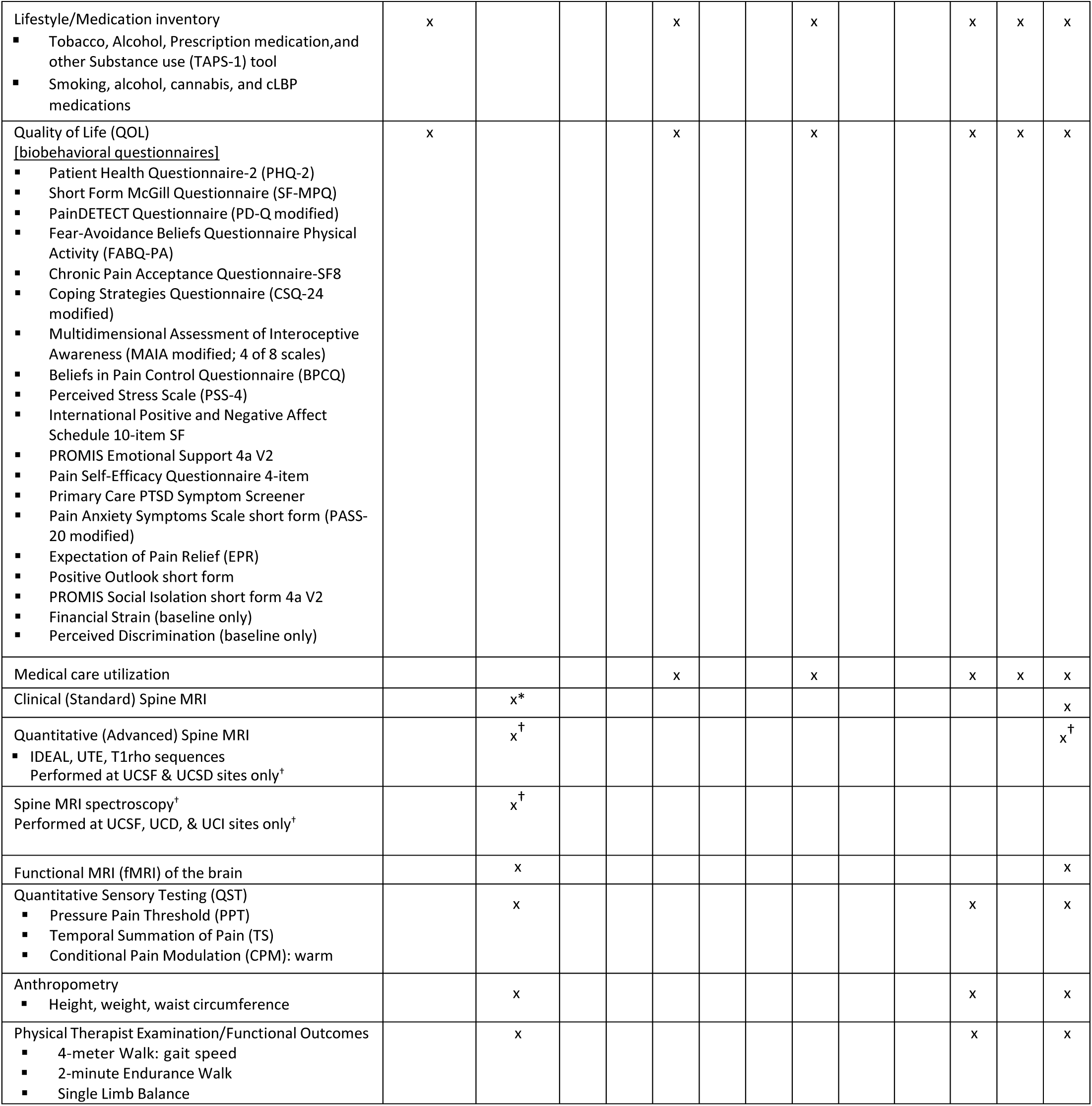
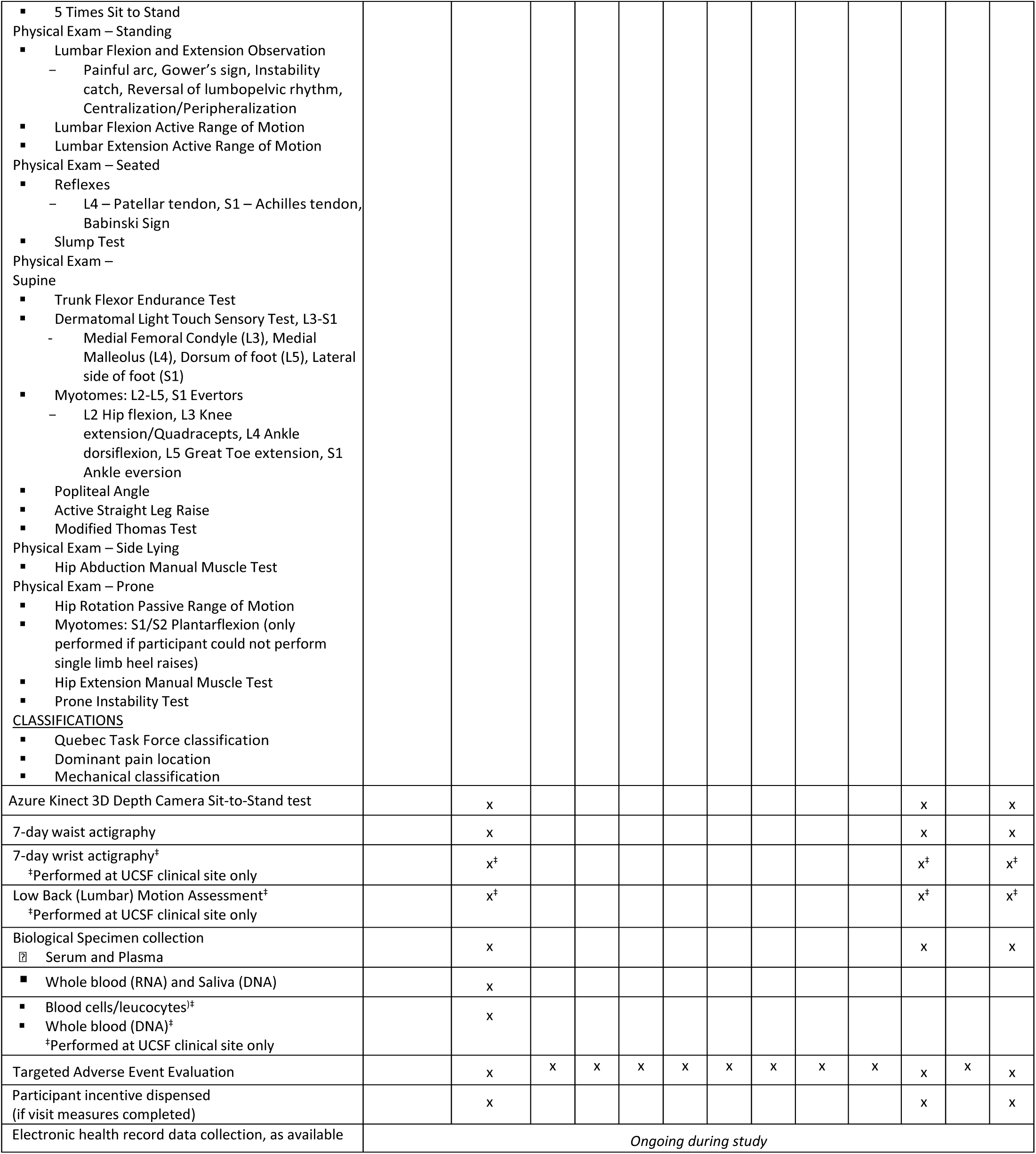
comeBACK Schedule of Evaluations and Visits.

**Table 3:**
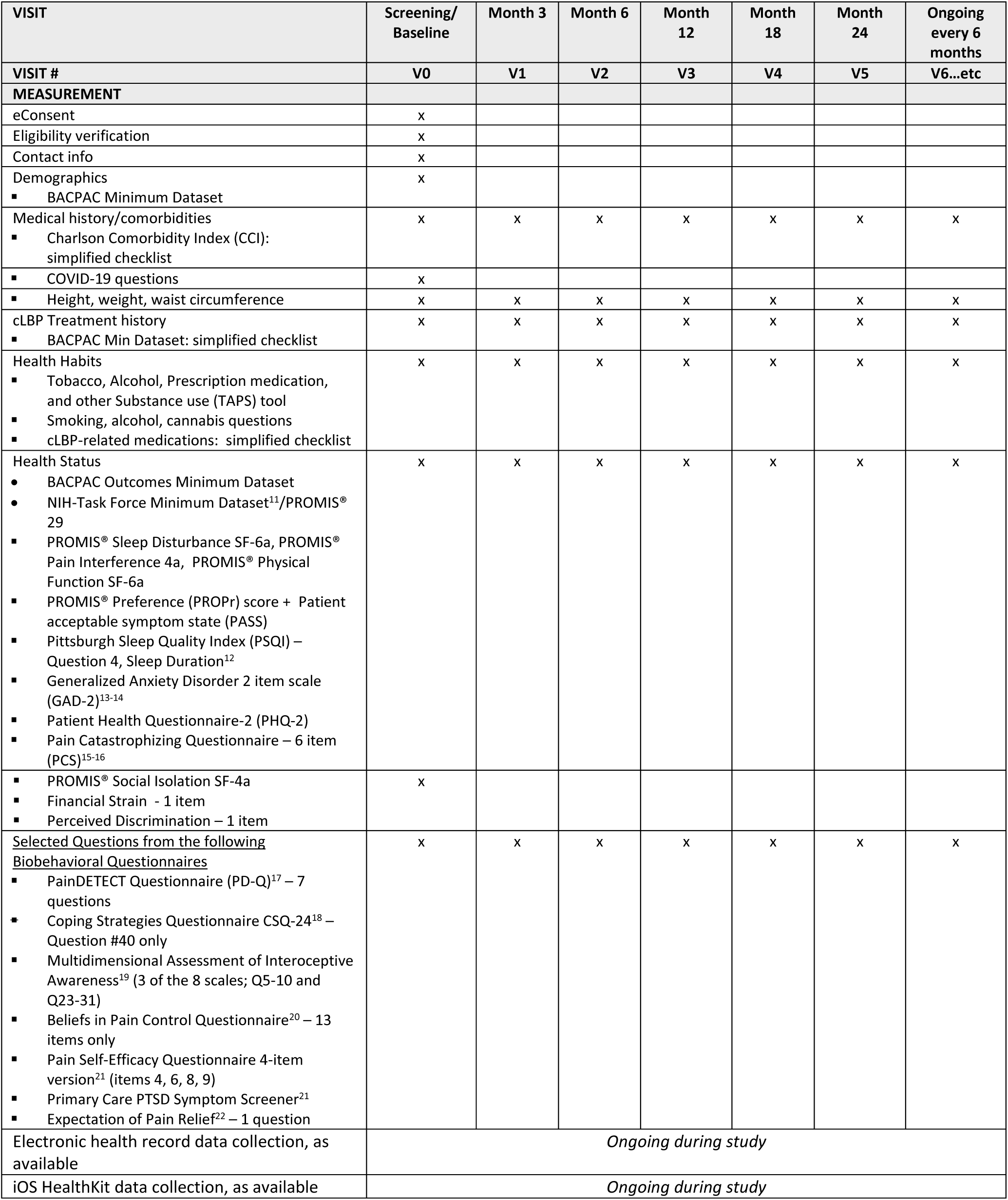
BACKHOME Schedule of Evaluations and Visits.

comeBACK IMAGING: The baseline and Month 24 or close-out visits included clinical and advanced magnetic resonance imaging (MRI) of the lumbar spine, consisting of T1 weighted and T2 weighted Fast Spin Echo Sequences (FSE) to assess disc, endplate, and bone marrow health, as well as quantitative lumbar spine MRI sequences (IDEAL, UTE, and T1rho) to assess disc quality (collagen and proteoglycan content), endplate morphology, spinal muscle fat content, and bone marrow fat content. Lumbar spine MR Spectroscopy (at baseline only) and fMRI of the brain (on a 3 Tesla scanner with sequences: Localizer Resting State fMRI, Field Map (GRE or SE -EPI based), T1w MPRAGE 1.0 mm, and T2w SPACE 1.0 mm SPACE 1.0 mm)) was also performed to assess what regions of the brain are involved in pain perception and how the brain responds. Advanced MRI and MRS studies are only performed at sites with the appropriate MRI machine available.

comeBACK CLINIC MEASURES: At baseline and each annual visit, comprehensive in-clinic testing includes: quantitative sensory function testing (QST), a psychophysical testing of cutaneous thermal sensibility, which examines the participant’s sensitivity to pressure, temporal summation of pain, and warm temperature; range of motion/strength tests and assessments of Quebec TaskForce classification, dominant pain location, and mechanical classification performed by a trained/study certified licensed Physical Therapist (PT) to ensure the necessary anatomical knowledge, biomechanical knowledge, and palpation skills to perform the tests successfully and reliably according to a standardized study protocol; functional outcome measurements (2 minute endurance test, 4 meter walk, 5 times sit to stand, and single limb balance test); a measure of waist actigraphy, where participants are instructed to wear an ActiGraph wearable device GT3X+ (Pensacola, FL, USA) at the hip level for 7 days; additionally at the UCSF clinical site, participants are also instructed to wear an ActiGraph device on their non-dominant wrist for 7 days to obtain wrist actigraphy; the Kinect 3D Depth Camera Sit to Stand test, a UCSF-developed test using a 3D Camera (Microsoft Azure Kinect, Redmond, WA, USA) to measure full body biomechanics during a standard sit-to-stand test with a marker-less single-camera motion capture system; a Low Back (Lumbar) Motion Assessment was also performed at the UCSF Clinical Site at the baseline visit only. This wearable motion sensor test is a short assessment of functional low back health that incorporates commercially available 9-axis Xsens MTw2 Inertial Measurement Unit Sensors (IMUs) and the Low Back Motion Assessment Pheno software (developed by The Ohio State University, Spine Research Institute (Columbus, OH, USA)) to assess the functional capabilities of the neuromusculoskeletal lumbar spine system.

### RECRUITMENT and RETENTION STRATEGIES

comeBACK participants receive up to $475 during study follow-up, for their time and effort to complete the study visits (i.e., an in-person visit may take 4-6 hours to complete). The participant incentives are offered as an amazon gift card, at the following timepoints: $150 gift card upon completion of baseline visit procedures, $150 gift card upon completion of the Month 3, 6, and 12 visit procedures, and $175 gift card upon completion of the Month 18 and 24 or close out visit procedures. Other retention strategies include: reminder emails and phone calls 1 week prior to an in-person visit, multiple follow-up emails if remote survey visits are not completed when received, phone calls to remind participants to complete remote survey visits or the Study Coordinator may also walk the participant through the survey over the phone, as needed, provision of parking at the clinical site, transportation to the site (e.g., Uber car service) if needed, provision of a lunch voucher for visits greater than 4 hours, and a snack bag after the phlebotomy measurement.

BACKHOME: Participants are not provided with any incentives for completing survey measures, but as a participant in BACKHOME, they will be eligible to be invited to future ancillary studies and interventions that are only available to participants in BACKHOME. For example, BACKHOME investigators are currently launching a remote ancillary study to collect outcomes via Apple Watches. Only BACKHOME participants are invited and will be eligible to receive Apple Watches as part of participation and study invitations will only be sent to eligible BACKHOME participants. Retention strategies include: multiple follow-up emails if remote survey visits are not completed when received and phone calls to remind participants to complete remote survey visits or the Study Coordinator may also walk the participant through the survey over the phone, as needed.

### DATA MONITIORING and TRAINING

In both studies, the UCSF CC is responsible for monitoring procedures related to study conduct and data collection/reporting to ensure the quality and integrity of the study data. For the comeBACK study, each clinical site investigator and coordinator were trained on the study protocol and procedures to ensure accurate and consistent study methods are used study-wide and throughout the entire study duration. Trainings include review of the protocol, operations manual, case report forms (CRFs), and data management procedures. At the start of the study, training was conducted at the comeBACK Investigators’ Meeting. When new clinical site staff were on-boarded, the UCSF CC conducted individual site trainings in-person or by web-meeting.

## STATISTICAL METHOD OVERVIEW

To analyze phenotypic traits, we plan to use a combination of traditional data analyses and deep learning methods, to define clinically useful cLBP phenotypes. To start, generalized linear mixed models will be used to assess associations of common data elements with measures of back pain and disability, which are to be assessed at baseline and follow-up visits; effects on trend will be captured by common data elements-time interactions.Clusters of statistically-significant common data elements will be considered as ‘phenotypes’. We will also use a combination of modern machine/deep learning techniques to associate phenotypes with outcomes of treatments that cohort subjects receive as a natural consequence of their cLBP care.

Sample Size in comeBACK: The research objectives of this project include defining personalized treatment goals, which constitute minimizing dysfunctions associated with most prioritized PROMIS-29 domains. Power calculations were performed for both cross-sectional analyses that assume the full study population and a longitudinal analysis that anticipates loss to follow-up over the two-year study period. The power calculations considered the ability to detect a significant Pearson correlation coefficient (*ρ*) between continuous risk factors and continuous primary outcomes (quantified by weak, moderate, and strong correlations ρ=0.1, 0.3, 0.5; Fisher’s Z-transformation), differences in continuous primary outcomes when stratified across four categorical risk factors equal prevalence meant to represent potential cLBP phenotypes (quantified by weak, moderate and strong effect sizes *f* = 0.1, 0.3, 0.5, 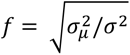; one-way analysis of variance [ANOVA]), and differences in predictive performance, specifically correlation, between pairs of supervised learning algorithms (quantified by pairwise differences among weak, moderate, and strong correlations, as defined above; Cohen’s q). Each power calculation assumes a two-sided test with a type-I error rate of 1% which is intended to provide a conservative estimate of power in acknowledgment of the fact that we do not account for multiplicity corrections.

The projected baseline recruitment for the comeBACK study was 400 participants (but we actually enrolled n=450). Based on prior clinical observational studies conducted by the UCSF CC, we anticipate 70-80% retention by the final two-year assessment yielding an effective sample size of at least n=400 participants for baseline cross-sectional analyses and n=315-320 for the two-year longitudinal analysis.

For the cross-sectional analysis with n=400, we would have 0.282, 0.998, and 1.000 power to detect a weak, moderate, and strong Pearson’s correlation coefficient, respectively, 0.160, 0.997 and 1.000 power to detect weak, moderate, and strong effect size for one-way ANOVA, respectively, and 0.645, 0.789, and 1.000 power to detect differences between weak and moderate, moderate and strong, and weak and strong Pearson’s correlation coefficients, respectively. For the longitudinal analysis with n=320, we have 0.215, 0.998, and 1.000 power to detect a weak, moderate, and strong Pearson’s correlation coefficient, respectively, 0.011, 0.985, 1.000 power to detect weak, moderate, and strong effect size for one-way ANOVA, respectively, and 0.523, 0.671, and 0.999 power to detect differences between weak and moderate, moderate and strong, and weak and strong Pearson’s correlation coefficients, respectively.

## SUMMARY

The University of California San Francisco’s (UCSF) Core Center for Patient-centric, Mechanistic Phenotyping in Chronic Low Back Pain (REACH), one of the three BACPAC Mechanistic Research Centers (MRCs) is conducting two large cohort translational research studies to help clarify biopsychosocial mechanisms of cLBP with the goal to provide a foundation to improve the evaluation of treatment effectiveness and to spur new avenues of therapeutic research for cLBP. The Longitudinal Clinical Cohort for Comprehensive Deep Phenotyping of Chronic Low-Back Pain (cLBP) Adults Study (comeBACK) is a multicenter in-person cohort study of 450 adults with chronic low back pain designed to perform comprehensive deep phenotyping in patients with cLBP. While, the eCohort of Chronic Low-Back Pain (cLBP) in Adults (BACKHOME) study is a site-less longitudinal observational e-cohort of approximately 3000 U.S. adults with cLBP. To our knowledge, this is the largest prospective remote registry of nationwide adults with chronic low back pain. These 2 pivotal studies, in conjunction with the ancillary studies being performed in both comeBACK and BACKHOME, and the other BACPAC-consortium research projects, we will be able to address a number of diagnostic and therapeutic issues in this complex and diverse patient population with cLBP.

## Data Availability

No study data was used for this manuscript. Data collection is currently ongoing and is not yet available.

## Acknowledgment

The Back Pain Consortium (BACPAC) Research Program is administered by the National Institute of Arthritis and Musculoskeletal and Skin Diseases (NIAMS).

## REFERENCES

1. Meucci RD, Fassa AG, Faria NM. Prevalence of chronic low back pain: systematic review. Rev Saude Publica. 2015;49:1. doi: 10.1590/S0034-8910.2015049005874. Epub 2015 Oct 20. PMID: 26487293; PMCID: PMC4603263.

2. GBD 2017 Disease and Injury Incidence and Prevalence Collaborators. Global, regional, and national incidence, prevalence, and years lived with disability for 354 diseases and injuries for 195 countries and territories, 1990-2017: a systematic analysis for the Global Burden of Disease Study 2017. Lancet. 2018 Nov 10;392(10159):1789–1858. doi: 10.1016/S0140-6736(18)32279-7. Epub 2018 Nov 8. Erratum in: Lancet. 2019 Jun 22;393(10190):e44. PMID: 30496104; PMCID: PMC6227754.

3. Manchikanti L, Singh V, Datta S, Cohen SP, Hirsch JA; American Society of Interventional Pain Physicians. Comprehensive review of epidemiology, scope, and impact of spinal pain. Pain Physician. 2009 Jul-Aug;12(4):E35–70. PMID: 19668291.

4. Esteban-Vasallo MD, Domínguez-Berjón MF, AstrayMochales J, Genova-Maleras R, Pérez-Sania A, Sánchez-Perruca L, et al. Prevalencia de enfermedades crónicas diagnosticadas en población inmigrante y autóctona. Gac Sanit. 2009;23(6):548–52. DOI:10.1590/S0213-91112009000600012

5. Melloh M, Röder C, Elfering A, Theis JC, Müller U, Staub LP, et al. Differences across health care systems in outcome and cost-utility of surgical and conservative treatment of chronic low back pain: a study protocol. BMC Musculoskelet Disord. 2008;9:81. DOI:10.1186/1471-2474-9-81

6. Liao ZT, Pan YF, Huang JL, Huang F, Chi WJ, Zhang KX, et al. An epidemiological survey of low back pain and axial spondyloarthritis in a Chinese Han population. Scand J Rheumatol. 2009;38(6):455–9. DOI:10.3109/03009740902978085

7. Loisel P, Lemaire J, Poitras S, Durand MJ, Champagne F, Stock S, et al. Cost-benefit and cost-effectiveness analysis of a disability prevention model for back pain management: a six year follow up study. Occup Environ Med. 2002;59(12):807–15. DOI:10.1136/oem.59.12.807

8. Lee SW, Patel J, Kim SY, Miranda-Comas G, Herrera J, Bartels MN. Use of Opioid Analgesics in Patients With Chronic Low Back Pain and Knee Osteoarthritis. Am J Phys Med Rehabil. 2019 Aug;98(8):e97–e98. doi: 10.1097/PHM.0000000000001109. PMID: 31318758.

9. Kamper SJ, Apeldoorn AT, Chiarotto A, Smeets RJ, Ostelo RW, Guzman J, van Tulder MW. Multidisciplinary biopsychosocial rehabilitation for chronic low back pain. Cochrane Database Syst Rev. 2014 Sep 2;2014(9):CD000963. doi: 10.1002/14651858.CD000963.pub3. PMID: 25180773; PMCID: PMC10945502.

10. Mauck MC, Lotz J, Psioda MA, Carey TS, Clauw DJ, Majumdar S, Marras WS, Vo N, Aylward A, Hoffmeyer A, Zheng P, Ivanova A, McCumber M, Carson C, Anstrom KJ, Bowden AE, Dalton D, Derr L, Dufour J, Fields AJ, Fritz J, Hassett AL, Harte SE, Hue TF, Krug R, Loggia ML, Mageswaran P, McLean SA, Mitchell UH, O’Neill C, Pedoia V, Quirk DA, Rhon DI, Rieke V, Shah L, Sowa G, Spiegel B, Wasan AD, Wey HM, LaVange L. The Back Pain Consortium (BACPAC) Research Program: Structure, Research Priorities, and Methods. Pain Med. 2023 Aug 4;24(Suppl 1):S3–S12. doi: 10.1093/pm/pnac202. PMID: 36622041; PMCID: PMC10403298.

11. Deyo RA, Dworkin SF, Amtmann D, Andersson G, Borenstein D, Carragee E, Carrino J, Chou R, Cook K, DeLitto A, Goertz C, Khalsa P, Loeser J, Mackey S, Panagis J, Rainville J, Tosteson T, Turk D, Von Korff M, Weiner DK. Focus article: report of the NIH Task Force on Research Standards for Chronic Low Back Pain. Eur Spine J. 2014 Oct;23(10):2028–45. doi: 10.1007/s00586-014-3540-3. PMID: 25212440.

12. Buysse DJ, Reynolds CF 3rd, Monk TH, Berman SR, Kupfer DJ. The Pittsburgh Sleep Quality Index: a new instrument for psychiatric practice and research. Psychiatry Res. 1989 May;28(2):193–213. doi: 10.1016/0165-1781(89)90047-4. PMID: 2748771.

13. Kroenke K, Spitzer RL, Williams JB, Monahan PO, Löwe B. Anxiety disorders in primary care: prevalence, impairment, comorbidity, and detection. Ann Intern Med. 2007 Mar 6;146(5):317–25. doi: 10.7326/0003-4819-146-5-200703060-00004. PMID: 17339617.

14. Kroenke K, Spitzer RL, Williams JB. The Patient Health Questionnaire-2: validity of a two-item depression screener. Med Care. 2003 Nov;41(11):1284–92. doi: 10.1097/01.MLR.0000093487.78664.3C. PMID: 14583691.

15. McWilliams LA, Kowal J, Wilson KG. Development and evaluation of short forms of the Pain Catastrophizing Scale and the Pain Self-efficacy Questionnaire. Eur J Pain. 2015 Oct;19(9):1342–9. doi: 10.1002/ejp.665. Epub 2015 Mar 11. PMID: 25766681.

16. Sullivan MJL, Bishop SR, Pivik J. The Pain Catastrophizing Scale: Development and validation. Psychological assessment. 1995;7(4):524–532.

17. Freynhagen R, Baron R, Gockel U, Tölle TR. painDETECT: a new screening questionnaire to identify neuropathic components in patients with back pain. Curr Med Res Opin. 2006 Oct;22(10):1911–20. doi: 10.1185/030079906X132488. PMID: 17022849.

18. Harland NJ, Georgieff K. Rehabilitation Psychology. 2003:48(4)296–300.

19. Mehling WE, Acree M, Stewart A, Silas J, Jones A. The Multidimensional Assessment of Interoceptive Awareness, Version 2 (MAIA-2). PLoS One. 2018 Dec 4;13(12):e0208034. doi: 10.1371/journal.pone.0208034. PMID: 30513087; PMCID: PMC6279042

20. Skevington SM. A standardised scale to measure beliefs about controlling pain (B.P.C.Q.): A preliminary study, Psychology and Health. 1990;4:3,221–232.

21. Nicholas M.K. Self-efficacy and chronic pain. Paper presented at the annual conference of the British Psychological Society. St. Andrews, 1989.

22. Prins A, Bovin MJ, Smolenski DJ, Marx BP, Kimerling R, Jenkins-Guarnieri MA, Kaloupek DG, Schnurr PP, Kaiser AP, Leyva YE, Tiet QQ. The Primary Care PTSD Screen for DSM-5 (PC-PTSD-5): Development and Evaluation Within a Veteran Primary Care Sample. J Gen Intern Med. 2016 Oct;31(10):1206–11. doi: 10.1007/s11606-016-3703-5. Epub 2016 May 11. PMID: 27170304; PMCID: PMC5023594.

23. Cormier S, Lavigne GL, Choinière M, Rainville P. Expectations predict chronic pain treatment outcomes. Pain. 2016 Feb;157(2):329–338. doi: 10.1097/j.pain.0000000000000379. PMID: 26447703.

